# Impact of extending lockdown on COVID-19 dynamic during 2021 in London: a mathematical modelling study

**DOI:** 10.1101/2025.04.02.25325123

**Authors:** Nacira Ramdani

**Affiliations:** Regional Veterinary Laboratory of El Oued, National Institute of Veterinary Medicine, Algeria

**Keywords:** COVID-19, epidemic simulation, lockdown, mathematical modelling, pandemic response, transmission dynamics

## Abstract

This study examines the potential effects of extending London’s third COVID-19 lockdown for an entire year (January–December 2021) using a deterministic SEAIRD compartmental model. The research compares real-world COVID-19 case data with simulated outcomes under prolonged lockdown conditions.

Key findings show that, in the real-world scenario, cases peaked on December 29, 2021, with over 36,000 daily infections, whereas in the simulation, strict lockdown measures led to near elimination of cases within 171 days. The simulated attack rate (0.085%) and cumulative incidence (7,517 cases) were significantly lower than real-world values (14.11% and 1.24 million cases, respectively).

The study highlights the effectiveness of strict public health interventions in controlling viral spread but also acknowledges the trade-offs, such as economic and social disruptions. These insights contribute to discussions on pandemic response strategies and the balance between containment measures and societal well-being.

**IMPACTS:**

- The study demonstrates how long-term lockdowns can effectively reduce the spread of contagious diseases, highlighting the importance of timely and strict public health interventions.
- It reveals the significant gap between real-world and simulated pandemic outcomes, showing that stricter measures could have substantially limited case numbers and fatalities.
- The research underscores the trade-offs of pandemic management, emphasizing the need for a balance between controlling infections and minimizing the social and economic impacts of containment measures.

## INTRODUCTION

The COVID-19 pandemic has left an indelible mark on societies worldwide, influencing not only public health but also the global economy. In London, the pandemic unfolded in several waves, with the first peak in April 2020 followed by subsequent surges in 2021, particularly with the rise of the Alpha and Delta variants. The highest peak was recorded in December 2021, with over 36,000 daily cases due to the Omicron variant. In response, London, like the rest of the UK, experienced multiple lockdowns, beginning with a national lockdown in March 2020, followed by a tiered system of regional restrictions. In January 2021 (Institute for Government, 2022), the city entered its third lockdown as cases surged. However, this lockdown, while critical in preventing further strain on the healthcare system, was lifted in March 2021, leading to a gradual reopening of society as the vaccine rollout began.

The peak in December 2021 demonstrated just how devastating and persistent the virus could be. It raises the question: What if the lockdown in London had been extended for a full year, potentially until December 2021? Could a prolonged period of strict measures have significantly altered the trajectory of the pandemic, reducing both the immediate and long-term damage caused by COVID-19? This analysis seeks to explore the potential impacts of extending the lockdown for a full year in London.

## MATERIALS AND METHODS

### Data

The London COVID-19 incidence data extracted from UK COVID-19 Dashboard (UK Health Security Agency [UKHSA], n.d.). The third lockdown in England began on 6 January 2021 and was lifted on 8 March 2021(Institute for Government, 2022). The population size of London in 2021 was assumed to be 8799800 (Population Data UK, n.d.).

### Mathematical models and assumptions

The SEAIRD compartmental model was used to simulate the dynamics of COVID-19, with the following compartments: S for susceptible individuals, E for exposed individuals who are infected but not yet infectious, I for symptomatic infectious individuals, A for asymptomatic infectious individuals, R for recovered individuals, and D for deceased individuals. Key assumptions included: transmission occurs from pre-symptomatic, symptomatic and asymptomatic individuals, with βs representing the transmission rate from symptomatic and pre-symptomatic individuals and βa from asymptomatic individuals. The incubation period is 5.2 days (He et al., 2020). Pre-symptomatic infection starts approximately 2 days before symptoms (He et al., 2020). Overall, the time spent in the E compartment is 3.6 days, with 80% progressing to pre-symptomatic and then to symptomatic infection and 20% to asymptomatic infection (Pollock & Lancaster, 2020). Symptomatic individuals either recover in 13.4 days (Byrne et al., 2020) or die in 17 days (Linton et al., 2020), with a 0.9% case fatality rate (Johns Hopkins University & Medicine, n.d.), while asymptomatic individuals recover in 8 days (Byrne et al., 2020). The model also accounts for the impact of lockdowns, where a scaling factor(α) reduces transmission rates. Population dynamics assumed that natural births and deaths were negligible, and that recovered individuals have permanent immunity due to short time frames.

The differential equations for the SEAIRD model are as follows:

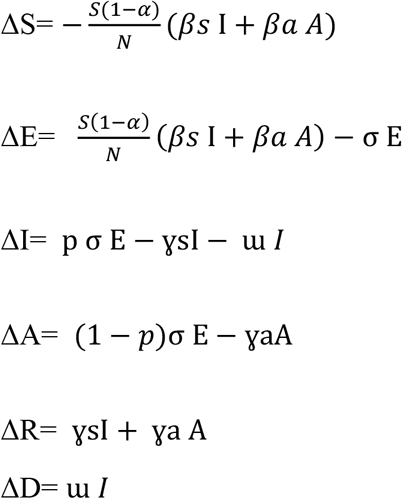

Where σ denotes the incubation rate, p denotes the proportion of exposed individuals that develop symptoms, ɣsI denotes the recovery rate from symptomatic infection, ɣadenotes recovery rate from asymptomatic infection and ɤ denotes the mortality rate.

This flow diagram represents the system of ordinary differential equations of the SEIDRD model

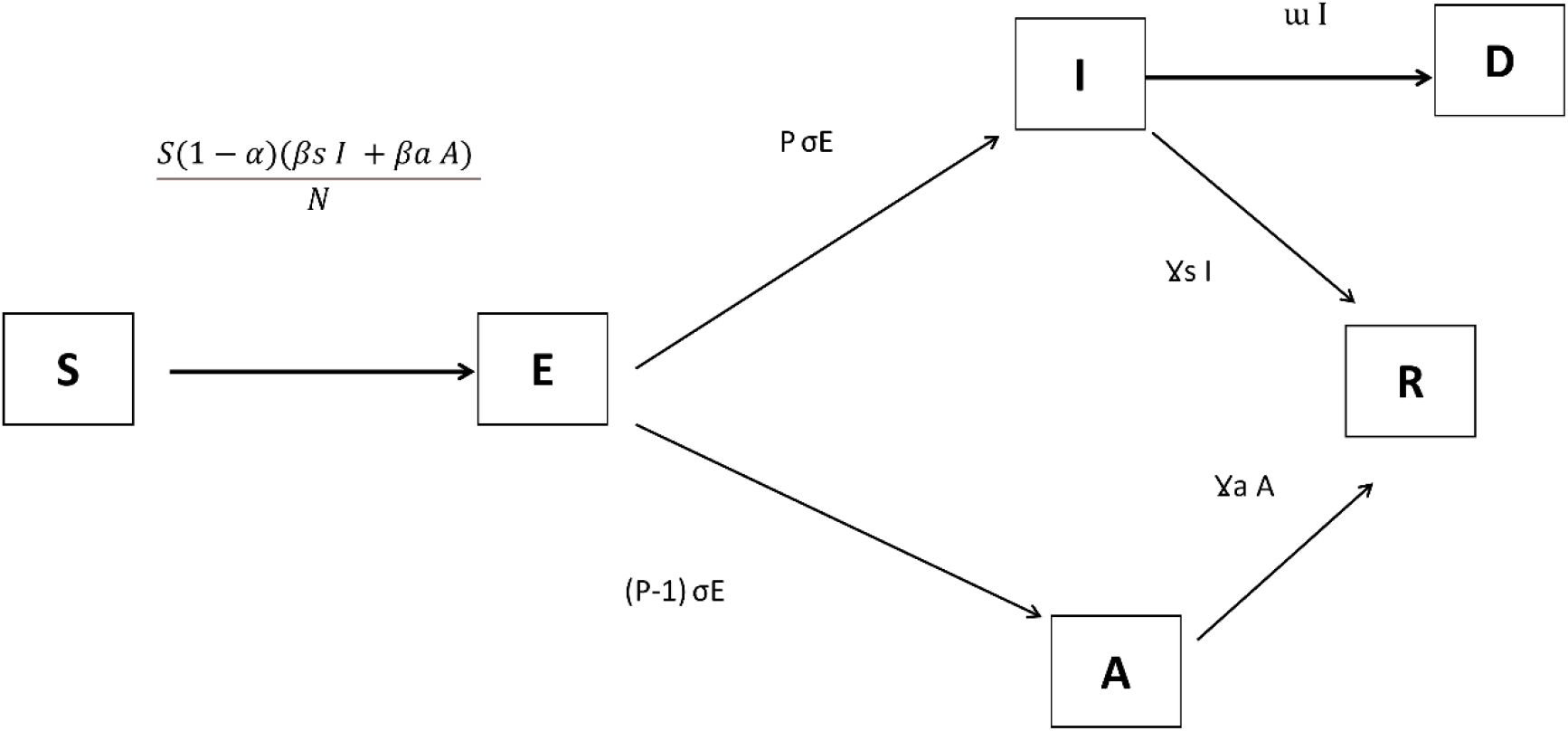

### Parameter estimation

To estimate the rates of transmission from symptomatic and asymptomatic infections *βs* and *βa* and the proportion of lockdown (α), the least sum of squares method was applied by fitting the deterministic SEIARD model to observed data during the third national lockdown in London from January 6 to March 7, 2021. These estimated parameters, combined with additional values from the literature, were subsequently used to simulate the dynamics of COVID-19 in London for the entirety of 2021. The initial values at time *t*_*0*_ correspond to those on the final day of the third lockdown (07/03/2021), derived from the simulated model calibrated to real-world data.

The analysis was performed using R, leveraging the ode function from the deSolve package alongside tools from the tidyverse. The resulting plots illustrate the simulated dynamics of COVID-19 in London under real measures implemented during 2021, including the impact of lockdown parameters on disease progression and outcomes.

The peaks of cases and peak prevalence were calculated to compare the timing and magnitude of the highest peaks during the lockdown and after its lifting in London in 2021. Cumulative incidence and attack rate were also computed.

## RESULTS

Based on the incidence data, the dynamics of COVID-19 in London reveal significant fluctuations in daily case numbers, reflecting the rise and fall of epidemic waves. The highest peak occurred on December 29, 2021, with 36,924 reported cases and a peak prevalence of 0.42%, indicating a substantial resurgence of the virus during the winter season. This peak aligns with a general upward trend observed throughout December 2021, where daily cases consistently exceeded 5000, reaching their zenith just before the year-end. Prior to this peak, smaller waves are evident, including one in January 2021, when cases ranged from 2,000 to nearly 18,000 daily, reflecting the impact of earlier stages of the pandemic. However, at the start of the third lockdown in the 6^th^ January, the epidemic of January started to decline gradually to reach 400 daily cases reflecting the gradual effectiveness of public health measures, such as lockdowns. Figure 1 illustrates the pattern of the disease dynamic.

**Figure 1:**
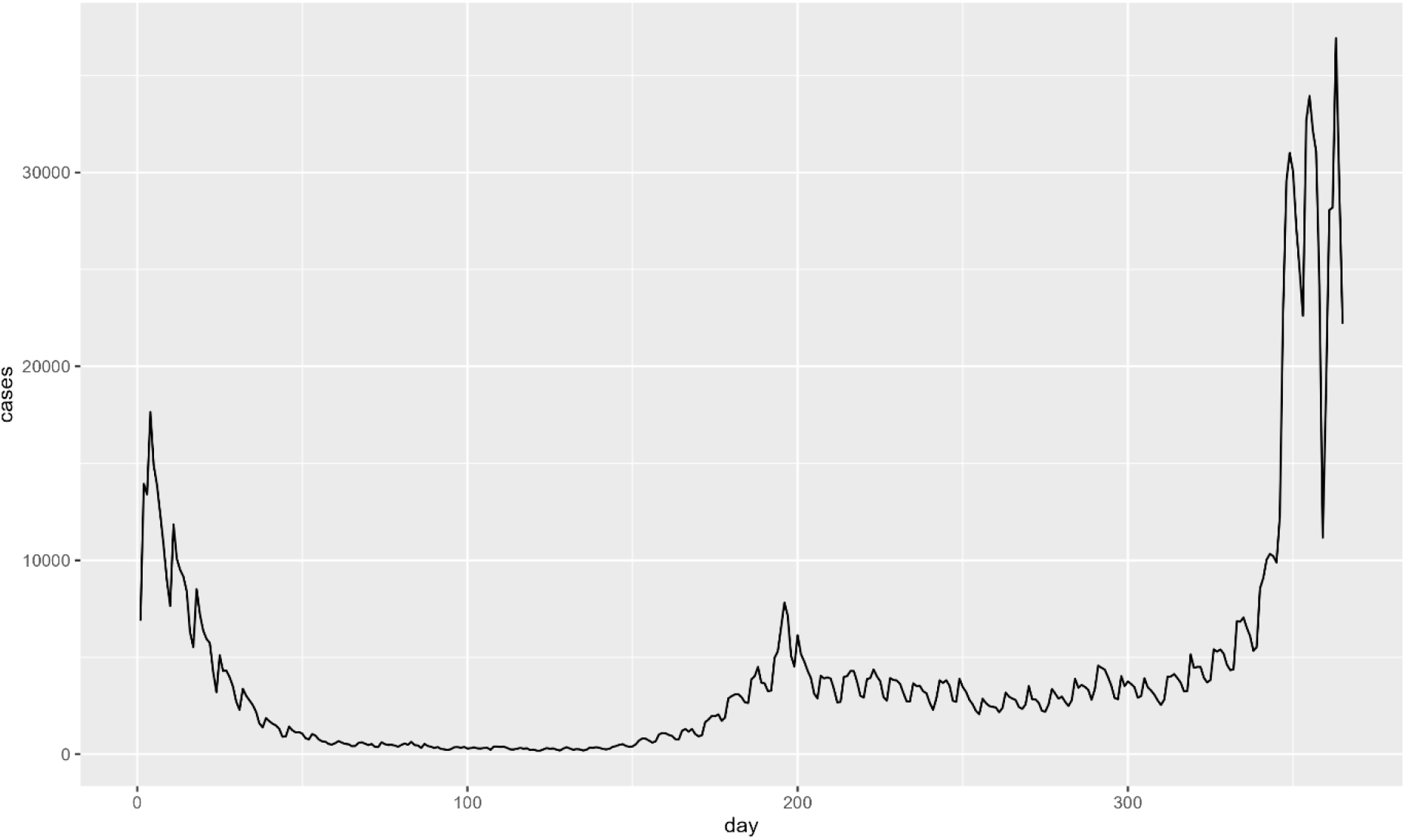
Dynamics of COVID-19 Daily Case Numbers in London, 2021

Under lockdown conditions, the estimated transmission rates of symptomatic and asymptomatic infections were found to be 0.4638491 and 0.01, respectively. The proportion of lockdown was 0.9629024.

Simulating the disease for a year with the lockdown parameters, the epidemic died out, the incidence decreased significantly until it reached one case within 171 days. The incidence peak was at time 1 from the starting point. However, the lowest point was at the last day of the year with less than one case (Figure 2).

**Figure 2:**
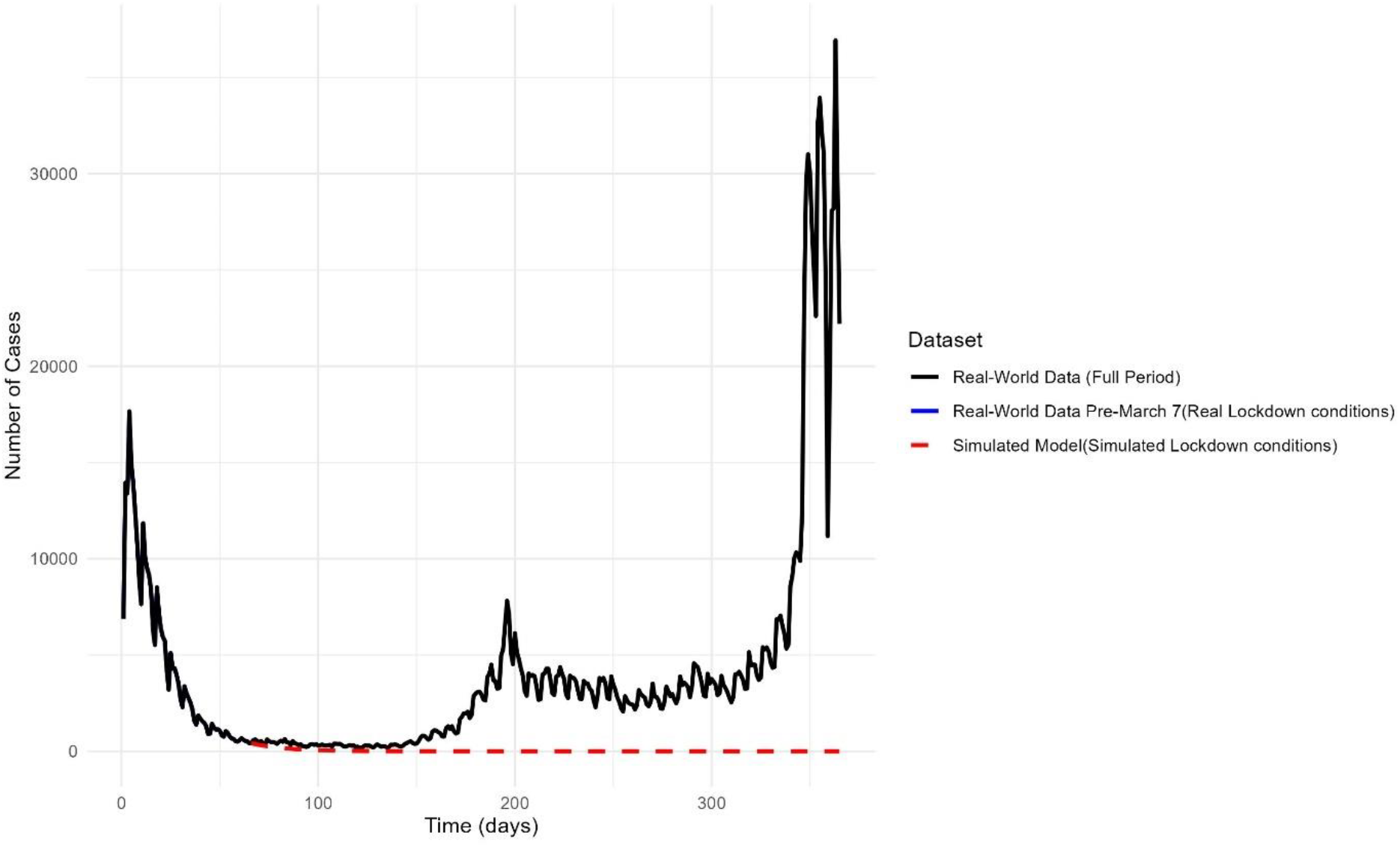
Comparison of simulated (Lockdown conditions) and real-world COVID-19 dynamics

**Figure 3:**
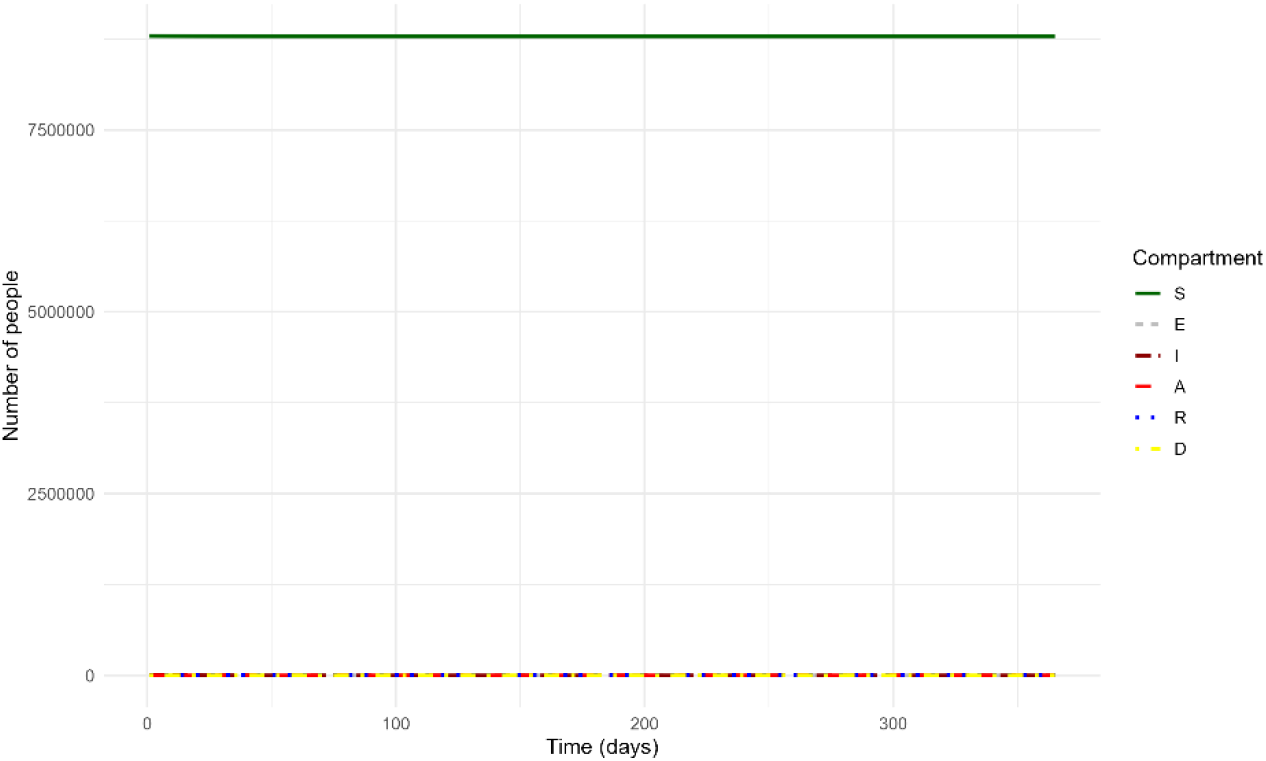
Dynamic of the Covid 19 under lockdown conditions in 2021 in London

The simulation of lockdown conditions for a year in London demonstrates that the epidemic dies out under such stringent measures, in contrast to the sustained transmission observed in real-world data from 2021. In the simulation, the peak occurred on 07-03-2021, representing the initial number of cases derived from the simulated model calibrated to real-world data, after which cases rapidly declined due to the lockdown. This contrasts sharply with the real-world peak on 29-12-2021, where the epidemic persisted and cases surged. The simulated cumulative incidence (7517.47 cases) and peak prevalence (0.0047%) were significantly lower than the real-world values of 1,241,653 cases and 0.42%, respectively, reflecting the effective containment of the virus in the simulated scenario. The attack rate (0.085%) in the simulation was also far below real-world levels (14.11%) (Table 1).

**Table 1.** Comparison of Simulated Lockdown Conditions and Real-World COVID-19 Data in London, 2021.

|  | Simulated (lockdown) | Real-world data |
| --- | --- | --- |
| Peak time | 07-03-2021 | 29-12-2021 |
| Cumulative reported incidence (I) | 7517.47 | 1241653 |
| Peak daily reported incidence (I) | 417.5068 | 36 924 |
| Peak prevalence (I) | 0.0047% | 0.42% |
| Attack rate (I) | 0.085% | 14.11% |

## DISCUSSION

The results highlight the stark differences between the dynamics of COVID-19 under simulated lockdown conditions and real-world outcomes in London during 2021. In the simulation, the epidemic quickly wanes under stringent lockdown measures, demonstrating the potential for such interventions to effectively curb viral transmission. The simulation’s outcome underscores the importance of early and sustained restrictions in suppressing outbreaks and reducing overall disease burden.

In contrast, the real-world data reflect the consequences of easing restrictions, with the epidemic persisting and peaking late in the year. This divergence between simulated and actual outcomes suggests that the relaxation of measures, coupled with other factors such as variant emergence, population mobility, and incomplete adherence to preventive measures, played a critical role in sustaining transmission.

The decision to lift the third lockdown in March 2021 was a pivotal moment in the UK’s response to the COVID-19 pandemic. While the lifting of restrictions allowed for economic recovery and social reopening, the resurgence of cases in late 2021 highlighted the persistent threat of the virus. This analysis revealed that a year-long lockdown, if implemented through December 2021, could have potentially reduced the devastating impact of the pandemic.

The potential benefits of a year-long lockdown include a more controlled spread of the virus, a stronger and more resilient healthcare system, and a faster vaccine rollout. However, these benefits would need to be weighed against the economic and social costs of prolonged restrictions. By analyzing these potential outcomes, we can gain valuable insights into the management of future pandemics and the complex trade-offs between public health measures and long-term societal recovery.

## Data Availability

COVID-19 case data for London is available on the UK Health Security Agency (UKHSA) dashboard

https://ukhsa-dashboard.data.gov.uk

## ACKNOWLEDGMENTS

We would like to thank Dr. Emma Davis for reviewing this work and providing valuable feedback.

## CONFLICT OF INTEREST STATEMENT

The author declares no competing interests.

## Notes

### Competing Interest Statement

The authors have declared no competing interest.

### Funding Statement

This study did not receive any funding

